# Influenza vaccine effectiveness against hospital-attended influenza infection in 2023/24 season in Hangzhou, China

**DOI:** 10.1101/2024.04.29.24306602

**Authors:** Hao Lei, Beidi Niu, Zhou Sun, Yaojing Wang, Xinren Che, Shengqiang Du, Yan Liu, Ke Zhang, Shi Zhao, Shigui Yang, Zhe Wang, Gang Zhao

## Abstract

**Background:** From 2020, influenza activities were largely affected by the coronavirus disease (COVID-19) pandemic at the global scale. The B/Yamagata lineage has become extinct since 2020, raising concerns regarding the quadrivalent influenza vaccine. Evaluating vaccine effectiveness (VE) against influenza infections is important to guide future influenza vaccine programs.

**Methods:** A test-negative case-control study was conducted in five tertiary hospitals in Hangzhou, the capital city of Zhejiang province, China. Hospital-attended patients aged >6 months who presented with influenza-like illness (ILI) from October 1, 2023 to March 31, 2024 were enrolled in this study. The VE was estimated using multivariate logistic regression models, adjusted by sex, age, influenza detection methods and influenza testing timing.

**Results:** In total, 157291 hospital-attended ILI participants were enrolled. 56704 (36%) were tested positive for influenza virus. The adjusted estimates of overall VE against any hospital-attended influenza infection was 48% (95% Confidence interval [CI]: 46%-51%). The overall VE of trivalent inactivated influenza vaccine (IIV3) was 59% (95% CI: 50%-66%), followed by trivalent live attenuated vaccine (LAIV3) (VE=53%, 95% CI: 42%-62%) and quadrivalent inactivated influenza vaccine (IIV4) (VE=47%, 95% CI: 45%-50%). IIV3 provided even much better protection against hospital-attended influenza B infection than IIV4 (VE=87% (95% CI: 81%-92%) for IIV3 versus VE=53%, 95% CI: 50%-57% for IIV4).

**Conclusions:** The influenza vaccine provided moderate protection against influenza infection in the 2023/24 season in Hangzhou, China, during a massive epidemic. The results supported the World Health Organization recommendation regarding the exclusion of B/Yamagata lineage antigen in quadrivalent influenza vaccines in 2023.

## Introduction

Influenza vaccination is the most effective way to prevent influenza infection [1]; however, the influenza vaccine effectiveness (VE) can vary considerably from year to year [2]. From 2020, the COVID-19 pandemic greatly disturbed influenza activities [3, 4]. In the 2023/24 season, a sharp increase in influenza activities was observed in China [5]. In addition, the B/Yamagata lineage became extinct in 2020, raising concerns regarding the quadrivalent influenza vaccine situation [6]. In September 2023, the World Health Organization (WHO) proposed that the inclusion of a B/ Yamagata lineage antigen in quadrivalent influenza vaccines is no longer warranted, and every effort should be made to exclude this component as soon as possible. [7]. Thus, evaluation of effectiveness of trivalent and quadrivalent influenza vaccines, especially the quadrivalent influenza vaccine, in the years without the B/Yamagata lineage has become significant. Globally, several studies have evaluated early or interim influenza VE after the COVID-19 pandemic [8–11]. However, to the best of our knowledge, no study had estimated the influenza VE for the entire flu season in 2023/24. As the timing of influenza vaccination plays a role in estimating its effectiveness [12], the influenza VE during the entire flu season may differ from the early or interim VE. To the best of our knowledge, no study has compared VE between trivalent and quadrivalent influenza vaccines in the 2023/24 season.

Since 2020, Zhejiang Province has provided free influenza vaccines to older adults aged >70 years [13]. This program provided an opportunity to evaluate the effectiveness of influenza vaccination policies in the real world. Recently, three types of influenza vaccines have been used in China: trivalent inactivated influenza vaccine (IIV3), quadrivalent inactivated influenza vaccine (IIV4), and trivalent live attenuated vaccine (LAIV3) [14]. For the trivalent influenza vaccine, in the 2023-2024 season, B/Victoria was used [15], and LAIV3 was used only in children aged 3-14 years.

Isolation of the virus and testing using real-time reverse-transcription polymerase chain reaction (RT-PCR) is adopted as the gold standard reference method for the diagnosis of the influenza [16]. And a test-negative case-control study design has been widely used to evaluate influenza VE [17]. However, small sample sizes and large ranges of the 95% confidence intervals (CIs) of the estimated VE have limited the robustness of the results, especially in China, with a relatively low influenza vaccination rate, which was approximately 1.5%-2.2% before the COVID-19 pandemic [18] and slight increased after the pandemic to 3.16% in 2020/21 season and 2.47% in 2021/22 season [19]. Although their sensitivity is relatively low, commercially available rapid antigen detection tests have the advantage of providing results much more quickly [16] and have been more widely used in hospitals in China. Thus, the large number of influenza-like illness (ILI) cases identified by antigen detection provided the possibility of conducting a robust VE evaluation and separate analysis of VE in subgroups in more detail. In addition, to the best of our knowledge, the impact of these two influenza detection methods on VE remains unknown.

In this study, we performed a test-negative case-control study to estimate the effectiveness of three different types of influenza vaccines against influenza infection in the 2023/24 season in Hangzhou city, China, during a period of massive influenza epidemic. We aimed to compare VE between three types of influenza vaccines (IIV3, IIV4, and LAIV3), and different age groups.

## Methods

### Study population

Samples were collected from five tertiary hospitals: First People’s Hospital of Hangzhou, Second People’s Hospital of Hangzhou, First People’s Hospital of Linping District, Zhejiang Xiaoshan Hospital, and First People’s Hospital of Xiaoshan District. Data were collected from October 1, 2023, to March 31, 2024, based on the seasonality of influenza epidemics in Zhejiang Province [20]. Regarding influenza detection, physicians were required to specify on the laboratory requisition form that the patient exhibited symptoms consistent with ILI. However, there is currently no standardized case definition of ILI [8]. Each month, individuals seeking medical attention for respiratory diseases with fever, accompanied by symptoms of cough and sore throat, and undergoing influenza testing, were enrolled in the study across the five healthcare institutions.

Eligible participants were patients with ILI who visited these five sentinel hospitals and were older than 6 months. Patients with an influenza testing interval exceeding 14 days were eligible for repeated inclusion in the study [2]. Basic demographic information, including sex and age, was collected for each patient with influenza testing, along with the detailed testing date, type of test method, and corresponding test results. Influenza testing methods included influenza nucleic acid detection by RT-PCR and influenza antigen detection. These two detection methods were analyzed individually to test the effect of the detection methods on the evaluation of VE. This study was approved by the Institutional Review Board and Human Research Ethics Committee of the School of Medicine of Zhejiang University (No. ZGL202404-1).

### Vaccination status

The virus strains recommended by the WHO in in the 2024 Northern Hemisphere influenza season were A/Victoria/4897/2022 (H1N1) pdm09-like virus, A/Darwin/9/2021 (H3N2)-like virus, and B/Austria/1359417/2021 (B/Victoria lineage)-like virus [15]. Patient vaccination information was obtained from the Zhejiang Province Electronic Medical System using the patient’s identity card (ID). This system comprehensively covers vaccination records for the majority of Hangzhou residents, as it was constructed to record COVID-19 vaccination status. Patients without vaccination records were excluded. Patient vaccination information from the Zhejiang Province Electronic Medical System included patient ID, vaccine type (IIV3, IIV4, or LAIV3), and vaccine date. This study specifically focused on the details of vaccine administration for 2022 and 2023. In accordance with Zhejiang Province’s vaccination policy, vaccination in the current season was defined as influenza vaccination from July 1, 2023, to March 31, 2024. Vaccination in the previous season was defined as influenza vaccination from July 1, 2022, to June 30, 2023. To explore the impact of previous vaccines on VE, the vaccination status of participants was categorized into four types: never vaccinated, vaccinated in both seasons, vaccinated only in the previous season, and vaccinated only in the current season.

Individuals were considered immunized if they had received an influenza vaccine for at leastLJ14 days before the specimen collection date [2]. Thus, influenza vaccines administered within 14 days before ILI onset were excluded from the analysis. The flu vaccine produced by manufacturers for children under 14 years of age requires two doses to be considered a complete vaccination.

### Statistical analysis

We pooled 6 months of data and compared characteristics of cases confirmed through influenza nucleic acid testing and influenza antigen testing with the characteristics of control cases that tested negative for influenza, stratified by virus type, vaccine type, and age group of patients. VE was calculated using a test-negative case-control design as (1 – OR) × 100, where OR is the odds ratio, which represents the odds of influenza virus infection among confirmed cases (positive test results) who received vaccination divided by the odds of vaccination among all influenza-negative cases (negative test controls). Adjusted ORs were determined using multivariate logistic regression models with adjustments for sex, age, influenza detection methods and month of influenza testing. Stratified VE estimates were computed based on the age group and influenza virus type. All statistical tests were conducted using a two-sided approach, and significance was defined as *P* < 0.05 or when the lower bound of the 95% CI for VE was greater than 0. Statistical analyses were performed using **R** statistical programming software version 4.2.1 (The R Project for Statistical Computing, Vienna, Austria).

### Sensitivity analysis

We conducted sensitivity analysis to assess the robustness of the estimated influenza VE. Firstly, we estimated the influenza VEs based on two influenza detection methods individually to test the impact of influenza detection methods. Second, to test the impact of influenza testing timing, we did not adjust the influenza testing timing or adjust the influenza testing data to estimate the VE individually.

## Results

### Participant characteristics

Data were collected from October 1, 2023, to March 31, 2024, involving a total of 157291 ILI patients tested for influenza (Table 1). Of which, 151,251 were tested by antigen and 6,040 were tested by nucleic acid (Table S1). The number of ILI patients tested for influenza peaked at week 53, 2023 (Figure 1). The dominant influenza subtype in the 2 years varied. Influenza A dominated in 2023, whereas influenza B dominated in 2024 (Figure 1). The overall influenza positivity rate was 36% in the ILI patients during the study period (Table 1), and the influenza positivity rates peaked at week 52 in 2023 at 49.9% (5,798/11,629) (Figure 1). By the end of this study, March 31, 2024, influenza activities also almost ended in Hangzhou (Figure 1). The weekly influenza positive rates of two detection methods were highly related (Pearson r=0.94, *P* < 0.001) (Figure S1). In the ILI patients, female was a little more than male (53.6% versus 46.4%). And people with the age between 18-59 years accounted for the highest proportion, comprising 50.4%. The overall influenza vaccine coverage rate was 7.1% (Table 1), with a coverage rate of 4.5% for positive cases and 8.6% for negative cases. The influenza vaccine coverage rate in the nucleic acid-tested patients was higher than that in the antigen-tested ILI patients (13.7% VS 6.8%) (Table S1), the main reason could be the much higher percentage of 70+ old people in the acid-tested participates than that in the antigen-tested patients (19.8% VS 1.14%), and higher vaccine coverage rate in old people (44.8% VS 21.7%) (Table S2). For these people who vaccinated in the 2023/24 season, the majority opted for IIV4, accounting for 86.3%, followed by IIV3 (8.8%) and LAIV3 (4.9%). And for these who vaccinated IIV3, most of them were older people with age more 70 years because the IIV3 influenza vaccines administered to older adults were free (Table S3). For people who were vaccinated in 2023-2024, the period between the influenza test date and the influenza vaccination date ranged from 15 to 249 days with a median of 97 days (Figure S2).

**Figure 1.**
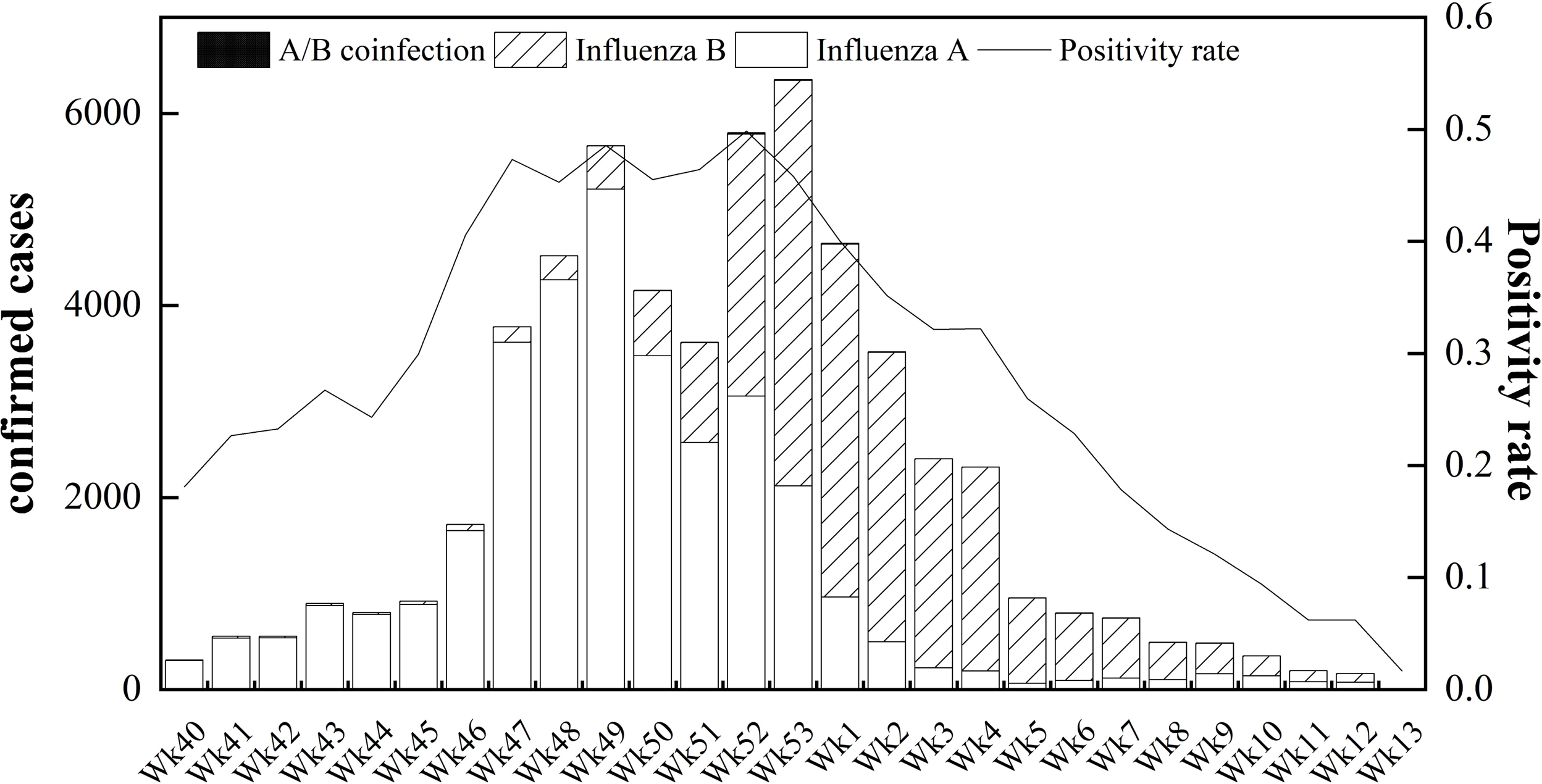
Weekly number of hospital-attended influenza infections and influenza positivity rates in five tertiary hospitals in Hangzhou.

**Table 1.**
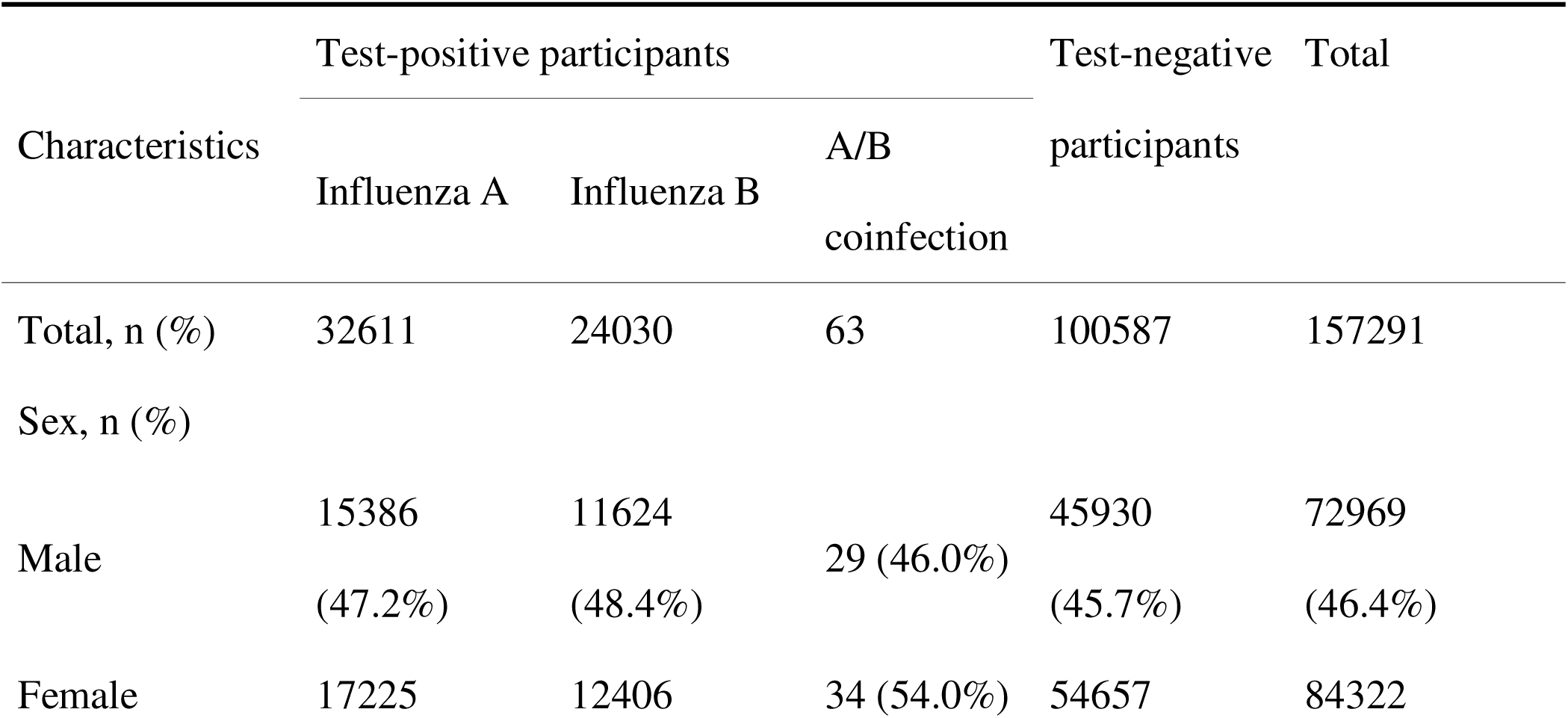
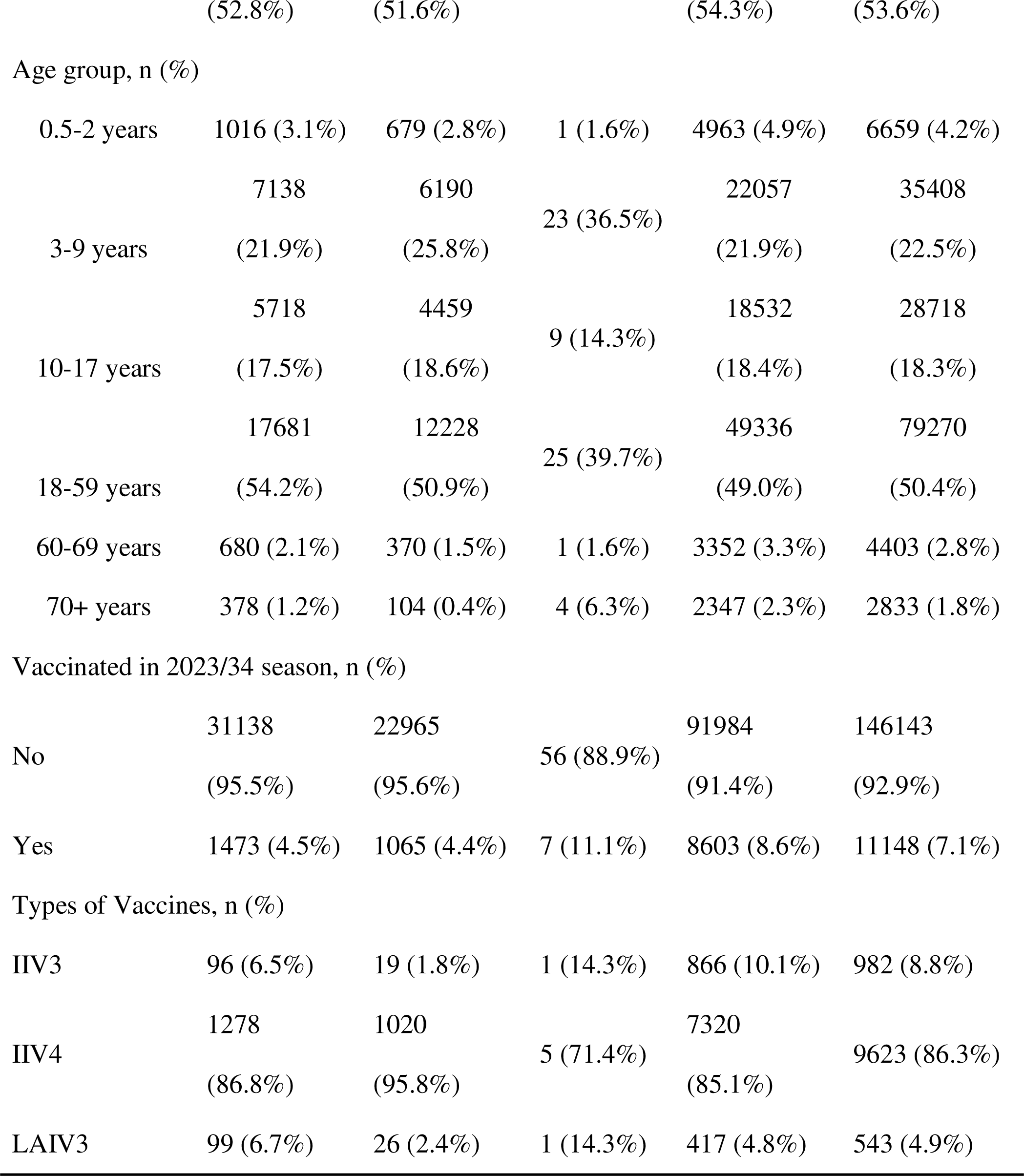
Demographic and clinical characteristics of participants who tested positive or negative for influenza virus.

### Overall VE

After adjustment for potential confounders, the estimated overall influenza VE against hospital-attended influenza infection in the 2023/24 season was 48% (95% confidence interval (CI): 46%-51%) (Figure 2). VE estimates were consistent between subjects using two influenza virus testing methods: 48% (95% CI: 46%-51%) for antigen-tested subjects, and 55% (95% CI: 44%-63%) for nucleic acid-tested subjects (Figure S3). However, because of the much larger number of patients detected by antigen testing, the estimated 95% CI of VE was much narrower. In addition, without adjusting the influenza testing timing or adjusted by the influenza testing date also had very limited impact on estimating the overall VEs (Figure S4). The overall VE of IIV3 was highest (VE=59%, 95% CI: 50%-66%), followed by LAIV3 (VE=53%, 95% CI: 42%-62%) and IIV4 (VE=47%, 95% CI: 45%-50%) (Figure 2). Adjusted VE was 65% for children aged 6 months to 2 years, 52% for people aged 18-59 years, 78% for old adults aged 60-69 years, and 28% for people aged above 70 (Figure 2). The VE against hospital-attended influenza B infection (VE=57%, 95% CI: 54%-60%) was much higher than that against influenza A infection (VE=38%, 95% CI: 34%-42%). (Figure 2).

**Figure 2.**
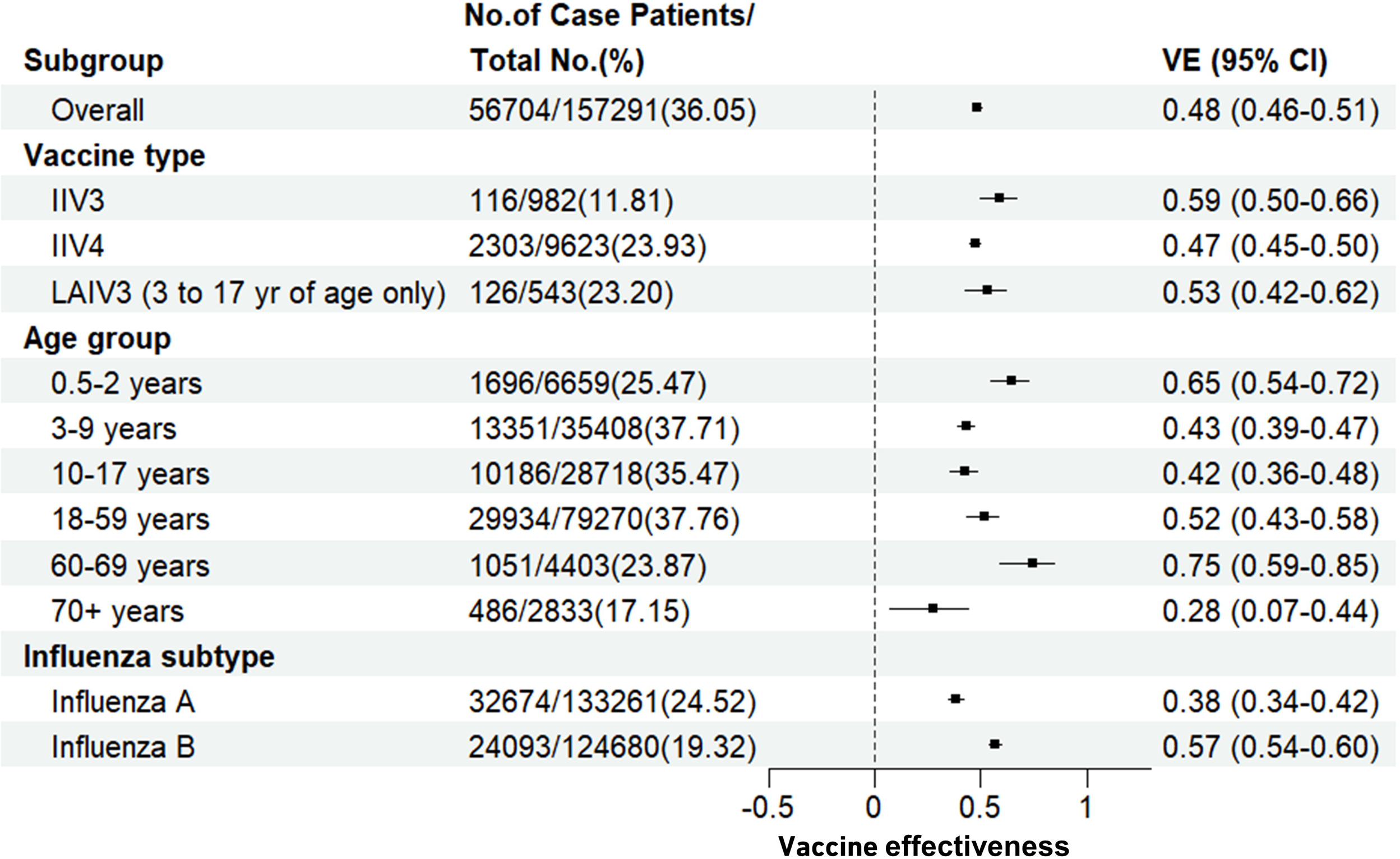
Adjusted estimates of influenza vaccine effectiveness (VE), overall and stratified to age groups, virus subtypes and vaccine types. Horizontal bars indicated 95% confidence intervals (CI).

### VE by influenza subtype, vaccine type and age group

Overall, influenza vaccine provided better protection against hospital-attended influenza B infection than against influenza A infection in 2023/24 season in Hangzhou. IIV3 provided much better protection than IIV4 against hospital-attended influenza B infection (Figure 3). The adjusted VE of IIV3 against influenza B infection was 87% (95% CI: 81%-92%), much higher than that of IIV4 against influenza B infection (VE=53%, 95% CI: 50%-57%). LAIV3 also provided high protection for children aged 3-17 years against influenza B infection (VE=79%, 95% CI: 69%-86%) (Figure 3). Though adjusted by age and testing timing, IIV4 provided best protection against hospital-attended influenza A infection, followed by LAIV3 and IIV3.

**Figure 3.**
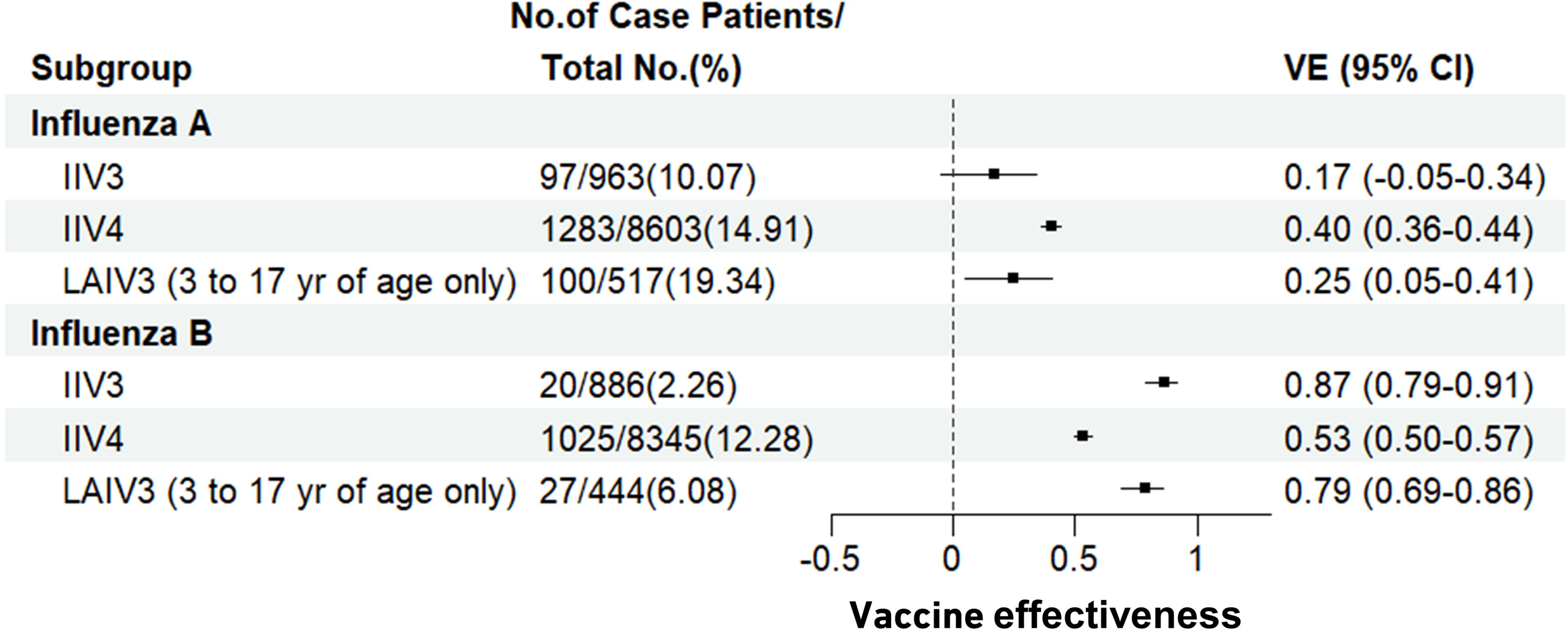
Adjusted VE against hospital-attended influenza A and B infection, stratified to vaccine types.

### VE for different vaccination seasons

VE against hospital-attended influenza infection was the highest for those vaccinated only in the current 2023/24 season (VE=52%, 95% CI=49%-55%) (Figure 4). Vaccination in the previous 2022/23 season could only provide low protection against hospital-attended influenza infection (VE=23%, 95% CI=18%-27%). However, vaccination in the previous 2022/23 season could still provide moderate protection against hospital-attended influenza B infection (VE=34%, 95% CI: 28%-40%). And vaccination in both seasons provided best protection against influenza B infection (VE=59%, 95% CI: 55%-63%). However, for influenza A, vaccination in the current season only provided best protection (VE=47%, 95% CI: 52%-60%) (Figure 4).

**Figure 4.**
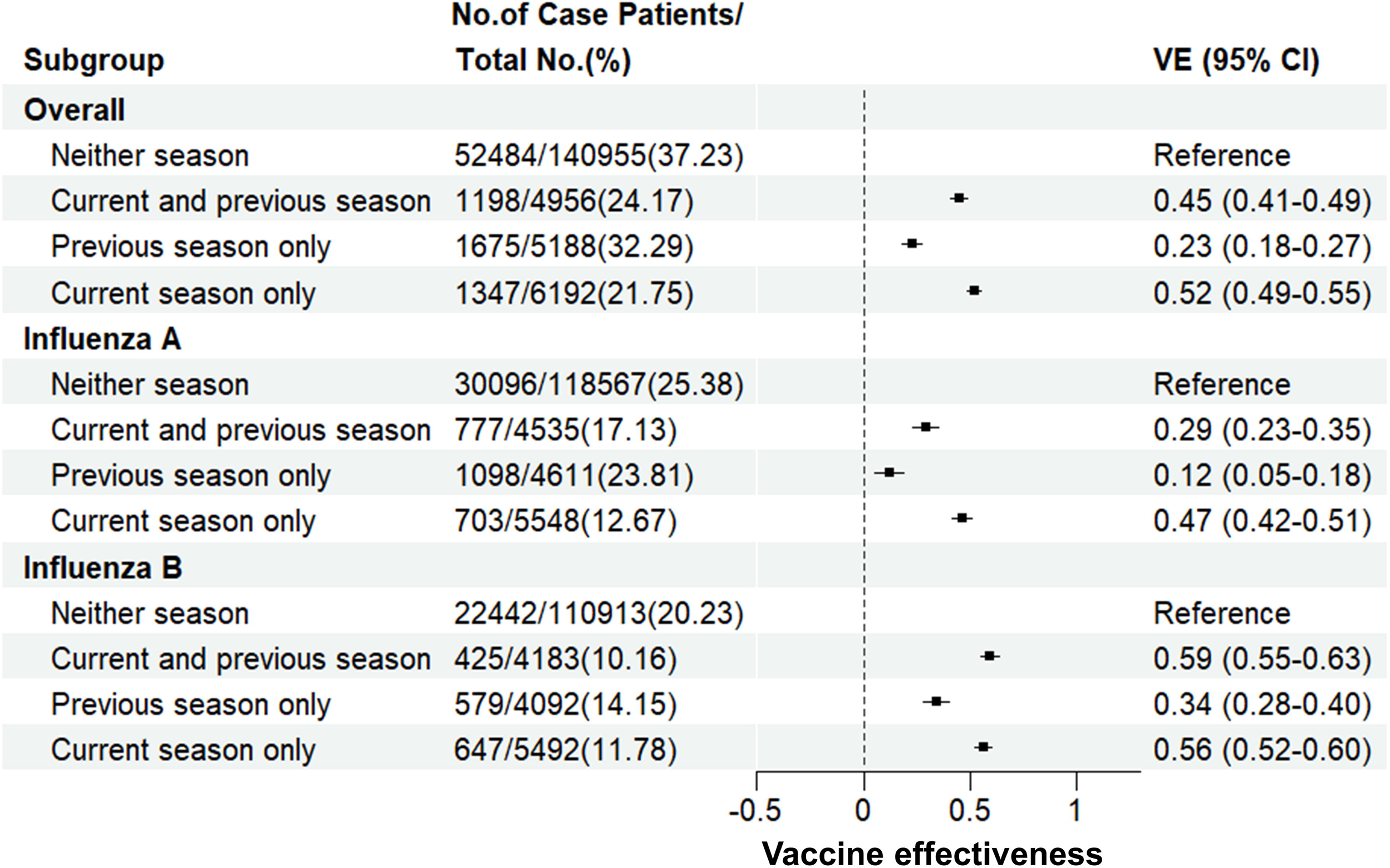
Adjusted VE against hospital-attended influenza A and B infection, stratified according to receipt of vaccines for the current season (2023/24) and previous season (2022/23).

## Discussion

Several studies have evaluated the effectiveness of influenza vaccines after the COVID-19 pandemic. In Denmark, VE was low at 24.8% in non-hospitalized patients aged 7–44 years in the 2021/22 season when there was a sharp increase in influenza detections [9]. In China, in the 2022/23 season when there was a slight increase in influenza infections, the estimated influenza VE was 56.3% against influenza infection in medically attended [10]. Several studies have estimated the effectiveness of early or interim influenza vaccines in the 2023-2024 season. In Canada, the estimated early influenza VE was 61% against influenza A H1N1, 49% against influenza A H3N2, and 75% against influenza B in the 2023/24 season [8]. In the Southern Hemisphere, the estimated interim influenza VE was 51.9% against influenza hospitalization in the 2023/24 flu season [11]. In California in the United States, the estimated interim influenza vaccine effectiveness against laboratory-confirmed influenza was 45% among persons aged ≥ 6 months [21]. However, to the best of our knowledge, no study has estimated the VE for the entire 2023/24 season. As the timing of influenza vaccination plays a role its effectiveness [12], the VE in the entire flu season may differ from the early or interim VE and would be more accurate in estimating the cost-effectiveness of the free influenza vaccination program. In this study, the estimated VE against influenza infection was 48% in the 2023/24 season in Hangzhou, China, which was close to the estimated interim VE in the Southern Hemisphere and US in the 2023/24 season [11, 21], And the estimated overall VE was robust to the influenza detection methods and influenza testing timing. The estimated VE in this study was much lower than the estimated early influenza VE in Canada [8], the timing of the studies may play a key role. However, similar to the study results in Canada, our study also found that influenza vaccines provided better protection against influenza B than influenza A in the 2023/24 season.

Before the COVID-19 pandemic, several studies also had evaluated the influenza VE in China in all age group in the year from 2013 to 2017 [23–25]. The estimated VE ranged from 5% (95% CI: −53%-41%) to 47% (95% CI: −20%-77%) in these studies. The observed variations could be due to the relatively small number of ILI patients enrolled in the studies, since the vaccine-effectiveness point estimates always have very wide confidence intervals in these studies. In addition, the influenza positivity rates in the ILI cases in these three studies ranged from 20% to 27% [23–25], much lower than the 36% influenza positivity rate in this study. This may suggest a relative higher VE during massive influenza epidemics, since the overall influenza VE was only 36% from a meta-analysis of the influenza VE before the pandemic in China [26].

The influenza vaccine coverage was 7.1% in the ILI patients in 2023/24 season in Hangzhou, which was much higher than the average influenza vaccine coverage rate in China, i.e., 2.47% in 2021/22 season [19], but was much lower than that in the United States [27]. The increased influenza vaccine coverage rate mainly contributed by the increased influenza vaccine coverage rate in the old people aged more than 70 years, which was 28%, much higher than these in other age groups. However, in Hangzhou in the 2023/24 season, we found that the majority opted IIV4, accounting for 86.3%, only 8.8% of ILI cases vaccinated IIV3. And for these who vaccinated IIV3, most of them were older people with age more 70 years because of the free influenza IIV3 program to them (Table S3). In this study, we found IIV3 provided better protection than IIV4 against influenza infection, especially against influenza B infection in the 2023/24 season (87% versus 52%). The main reason could be that B/Victoria used influenza vaccine components in trivalent influenza vaccine, since B/Yamagata lineage has become extinct [6]. Our results supported the report from WHO about the exclusion of B/Yamagata lineage antigen in quadrivalent influenza vaccines as soon as possible [7]. On March 14, 2024, the Centers for Diseases Control and Prevention of the United States. also announced the transition from quadrivalent to trivalent flu vaccines for the 2024-2025 season [22]. Since IIV3 is much cheaper than IIV4, but could provide better protection, the cost-effectiveness of IIV3 could also be much higher than IIV4.

The findings of this study have the following limitations. First, there was a preference among the population for antigen tests over nucleic acid tests, resulting in a smaller patient sample size for the latter. This has led to an inability to produce highly statistically reliable VE estimates in multiple subgroup analyses. Consequently, VE estimates for patients diagnosed using nucleic acid tests should be interpreted with caution. Second, we did not acquire data on the genetic or antigenic characteristics of the circulating A(H1N1) pdm09 or A(H3N2) strains in Hangzhou. Future research should focus on the antigenicity and genetic characteristics of these viruses to conduct more in-depth studies. Third, there are limitations associated with the use of routine health data collected by hospitals. For instance, the absence of symptomatic status data precludes the determination of whether cases are ILIs. Additionally, because of the reliance on specimen collection dates as proxies for symptom onset dates, the actual date of illness onset for most individuals was likely earlier. This may result in the misclassification of individuals as either influenza-negative or positive. For example, if the vaccination date and the onset of symptoms are close, but there is a long interval before testing. Lastly, other patient information that may influence the effectiveness of influenza vaccines, was not available, such as chronic diseases related to severe influenza. These chronic diseases include cardiovascular diseases, chronic respiratory diseases, neurological disorders, and diabetes [2].

## Conclusion

The influenza vaccine provided moderate protection against influenza infection during the massive epidemic in Hangzhou in the 2023/24 season. The large number of patients with an ILI detected by antigen testing made it possible to conduct a detailed separate analysis of influenza VE, such as by vaccine type, age group of patients, and influenza subtype. IIV3 provided better protection against hospital-attended influenza infection, especially influenza B. Thus, our study results support the WHO recommendation regarding the exclusion of B/Yamagata lineage antigens in quadrivalent influenza vaccines in September 2023.

## Acknowledgments

HL and BN conceived and designed the study. ZS, ZW, KZ, XC and YL collected data. HL and BN wrote the drafts of the manuscript. BN cleaned and analyzed the data. GZ, ZW and HL supervised the study. YW, SZ and SY commented on and revised drafts of the manuscript. HL interpreted the findings. All authors read and approved the final report.

## Funding

This study was supported by the innovation group on intelligent response to infectious diseases and public health emergencies of School of Public Health, Zhejiang University.

## Competing interests

All authors declare no competing interests.

## Data Availability

The data that support the findings of this study are available from the corresponding author upon reasonable request.

## Notes

### Competing Interest Statement

The authors have declared no competing interest.

### Author Declarations

the Institutional Review Board and Human Research Ethics Committee of the School of Medicine of Zhejiang University (No. ZGL202404-1) gave ethical approval for this work

## References

1. Paules C, Subbarao K. Influenza. Lancet, 2017 ; 390(10095):697–708.

2. Jackson ML, Chung JR, Jackson LA, et al. Influenza vaccine effectiveness in the United States during the 2015–2016 season. New England Journal of Medicine, 2017 ; 377(6): 534–543.

3. Lei H, Xu M, Wang X, et al. Nonpharmaceutical interventions used to control COVID-19 reduced seasonal influenza transmission in China. The Journal of infectious diseases, 2020 ; 222(11): 1780–1783.

4. Lei H, Yang L, Yang M, et al. Quantifying the rebound of influenza epidemics after the adjustment of zero-COVID policy in China. PNAS nexus, 2023; 2(5): pgad152.

5. Chinese National Influenza Center. Influenza Weekly Report. Week 3, 2024. Available at: https://ivdc.chinacdc.cn/cnic/en/Surveillance/WeeklyReport/202401/t20240125_272252.htm. Accessed 31 January 2024.

6. Monto A S, Zambon M, Weir J P. The End of B/Yamagata Influenza Transmission-Transitioning from Quadrivalent Vaccines. The New England journal of medicine, 2024 ; 390(14):1256–1258.

7. WHO. Recommended composition of influenza virus vaccines for use in the 2024 southern hemisphere influenza season. Wkly Epidemiol Rec 2023; 98: 521–33. Available at: https://iris.who.int/bitstream/handle/10665/373645/WER9843enfre.pdf. Accessed 31 January 2024.

8. Smolarchuk C, Ickert C, Zelyas N, et al. Early influenza vaccine effectiveness estimates using routinely collected data, Alberta, Canada, 2023/24 season [J]. Euro Surveill, 2024 ; 29(2).

9. Emborg HD, Vestergaard L S, Botnen A B, et al. A late sharp increase in influenza detections and low interim vaccine effectiveness against the circulating A (H3N2) strain, Denmark, 2021/22 influenza season up to 25 March 2022[J]. Eurosurveillance, 2022 ; 27(15): 2200278.

10. Su Y, Guo Z, Gu X, et al. Influenza vaccine effectiveness against influenza A during the delayed 2022/23 epidemic in Shihezi, China[J]. Vaccine, 2023 ; 41(39): 5683–5686.

11. Fowlkes A L. Interim Effectiveness Estimates of 2023 Southern Hemisphere Influenza Vaccines in Preventing Influenza-Associated Hospitalizations—REVELAC–i Network, March–July 2023[J]. MMWR. Morbidity and Mortality Weekly Report, 2023 ; 72.

12. Worsham C M, Bray C F, Jena A B. Optimal timing of influenza vaccination in young children: population based cohort stud. BMJ, 2024 ; 384.

13. Zhang X, Shen P, Liu J, et al. Evaluating the effectiveness and cost-effectiveness of free influenza vaccination policy for older adults in Yinzhou, China: Study protocol of a real-world analyses. Vaccine, 2023 ; 41(34): 5045–5052.

14. TWGIV, National Immunization Advisory Committee, Technical Working Group. Technical guidelines for seasonal influenza vaccination in China (2023-2024). Zhonghua liu xing bing xue za zhi, 2023 ; 44(10): 1507–1530.

15. World Health Organization. Recommended composition of influenza virus vaccines for use in the 2024 northern hemisphere influenza season. 2023. Available at: https://www.who.int/publications/m/item/recommended-composition-of-influenza-virus-vaccines-for-use-in-the-2023-2024-northern-hemisphere-influenza-season. Accessed 31 January 2024.

16. Ghebremedhin B, Engelmann I, König W, et al. Comparison of the performance of the rapid antigen detection actim Influenza A&B test and RT-PCR in different respiratory specimens. Journal of medical microbiology, 2009 ; 58(3): 365–370.

17. Fukushima W, Hirota Y. Basic principles of test-negative design in evaluating influenza vaccine effectiveness. Vaccine. 2017 ; 35(36):4796–4800.

18. Yang J, Atkins KE, Feng L, et al. Seasonal influenza vaccination in China: Landscape of diverse regional reimbursement policy, and budget impact analysis. Vaccine. 2016 ; 34(47):5724–5735.

19. Zhao H, Peng Z, Ni Z, et al. Investigation on influenza vaccination policy and vaccination situation during the influenzaseasons of 2020-2021 and 2021-2022 in China. Chinese Journal of Preventive Medicine, 2022 ; 56(11): 1560–1564.

20. Lei H, Yang L, Wang G, et al. Transmission patterns of seasonal influenza in China between 2010 and 2018. Viruses, 2022 ; 14(9): 2063.

21. Zhu S. Interim Influenza Vaccine Effectiveness Against Laboratory-Confirmed Influenza—California, October 2023–January 2024[J]. MMWR. Morbidity and Mortality Weekly Report, 2024 ; 73.

22. Centers for Diseases Control and Prevention. Seasonal Flu Vaccines. Available at https://www.cdc.gov/flu/prevent/flushot.htm. Accessed 26 April 2024.

23. Qin Y, Zhang Y, Wu P, et al. Influenza vaccine effectiveness in preventing hospitalization among Beijing residents in China, 2013–15[J]. Vaccine, 2016 ; 34(20): 2329–2333.

24. Zhang L, Pan Y, Hackert V, et al. The 2015–2016 influenza epidemic in Beijing, China: Unlike elsewhere, circulation of influenza A (H3N2) with moderate vaccine effectiveness[J]. Vaccine, 2018 ; 36(33): 4993–5001.

25. Wu S, Pan Y, Zhang X, et al. Influenza vaccine effectiveness in preventing laboratory-confirmed influenza in outpatient settings: a test-negative case-control study in Beijing, China, 2016/17 season[J]. Vaccine, 2018 ; 36(38): 5774–5780.

26. Yang X, Zhao H, Li Z, et al. Influenza vaccine effectiveness in mainland China: a systematic review and meta-analysis[J]. Vaccines, 2021 ; 9(2): 79.

27. Centers for Diseases Control and Prevention. Flu Vaccination Coverage, United States, 2022–23 Influenza Season. Available at https://www.cdc.gov/flu/fluvaxview/coverage-2223estimates.htm Accessed 28 April 2024.

