## Supplementary Material for "Influenza vaccine effectiveness against hospital-attended influenza infection in 2023/24 season in Hangzhou, China"

**Supplementary materials**

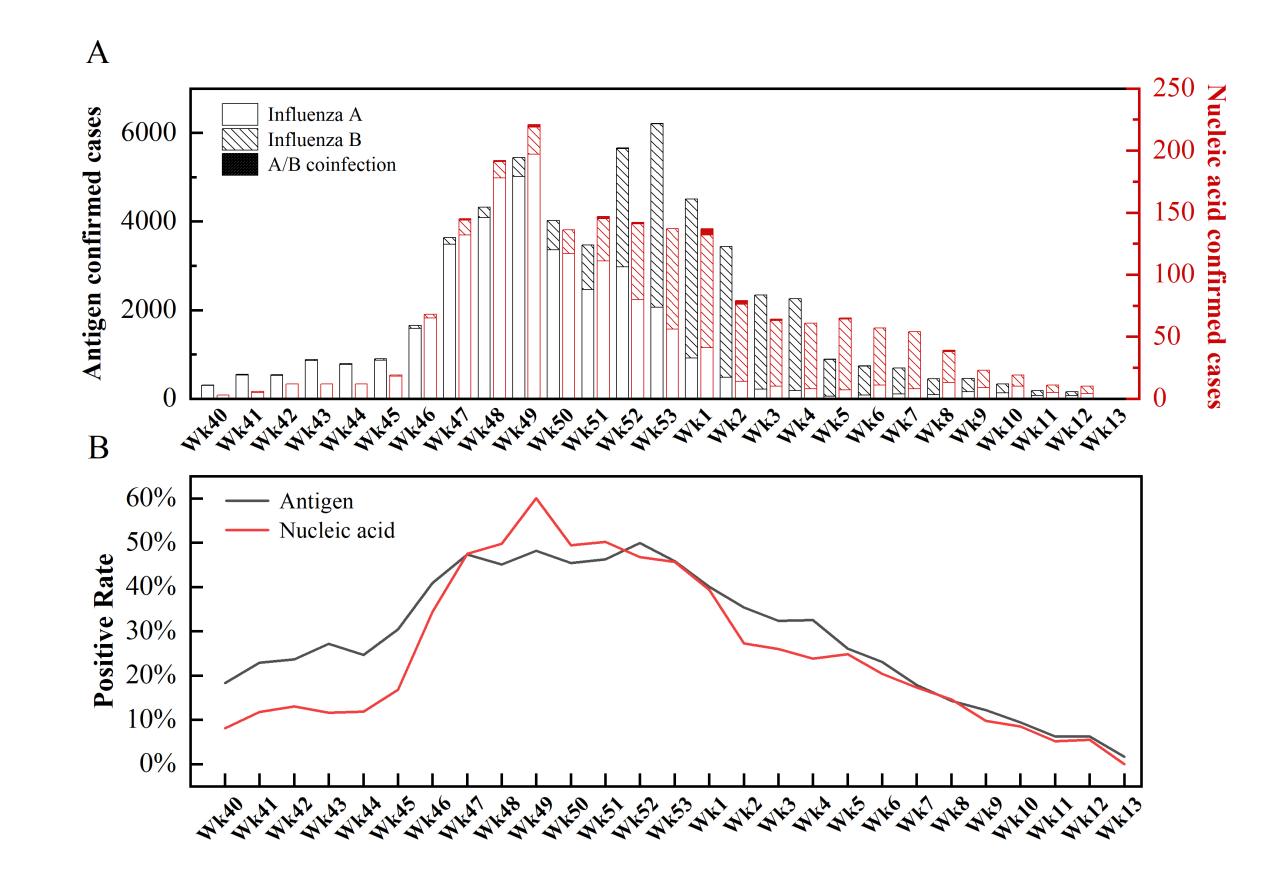

**Figure S1**. Weekly influenza activities detected by antigen and nucleic acid respectively in five hospitals in Hangzhou. (A) number of influenza A, B and co-infection cases. (B) weekly influenza positivity rates.

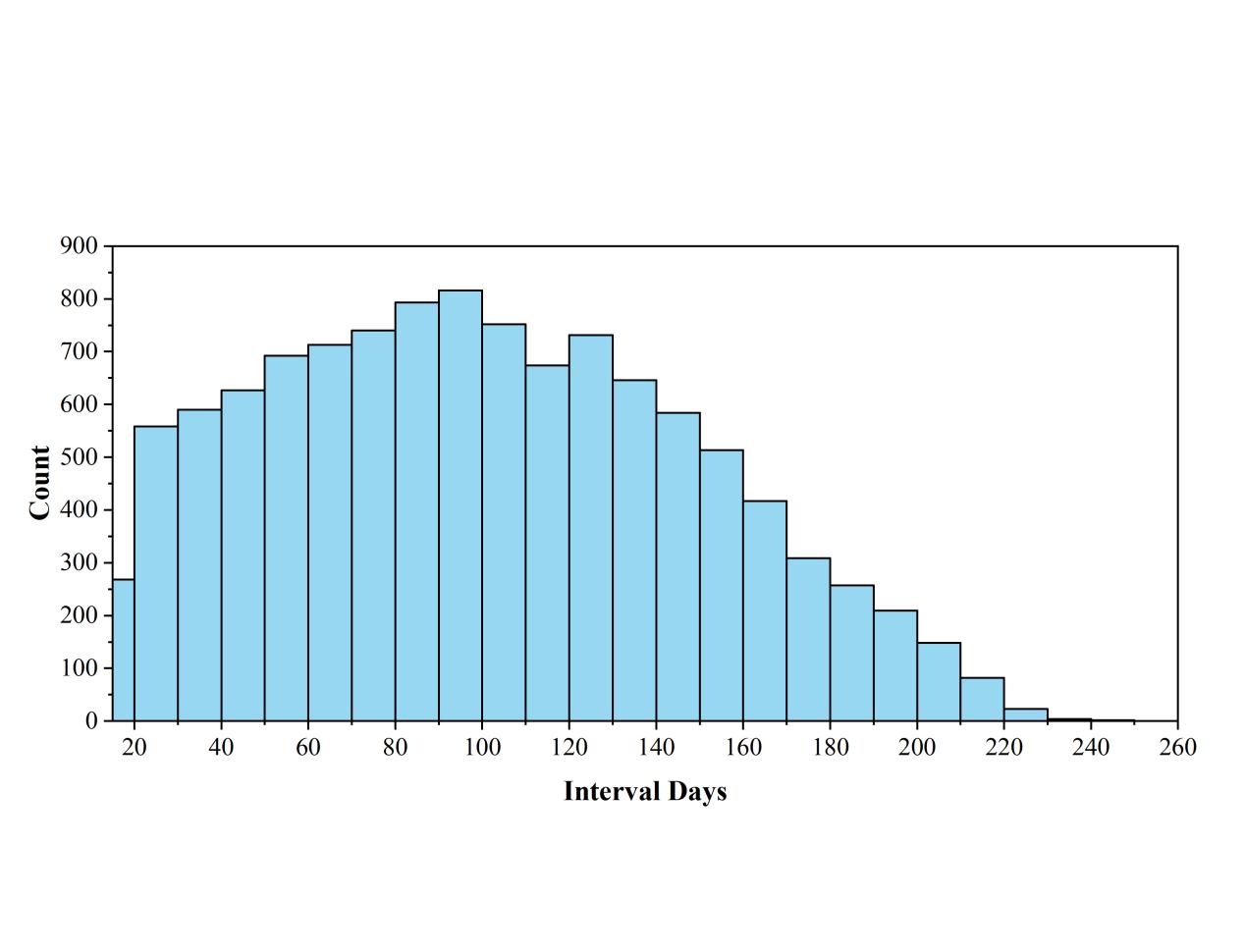

**Supplementary Figure S2**. Distribution of the interval between vaccination date and influenza test date.

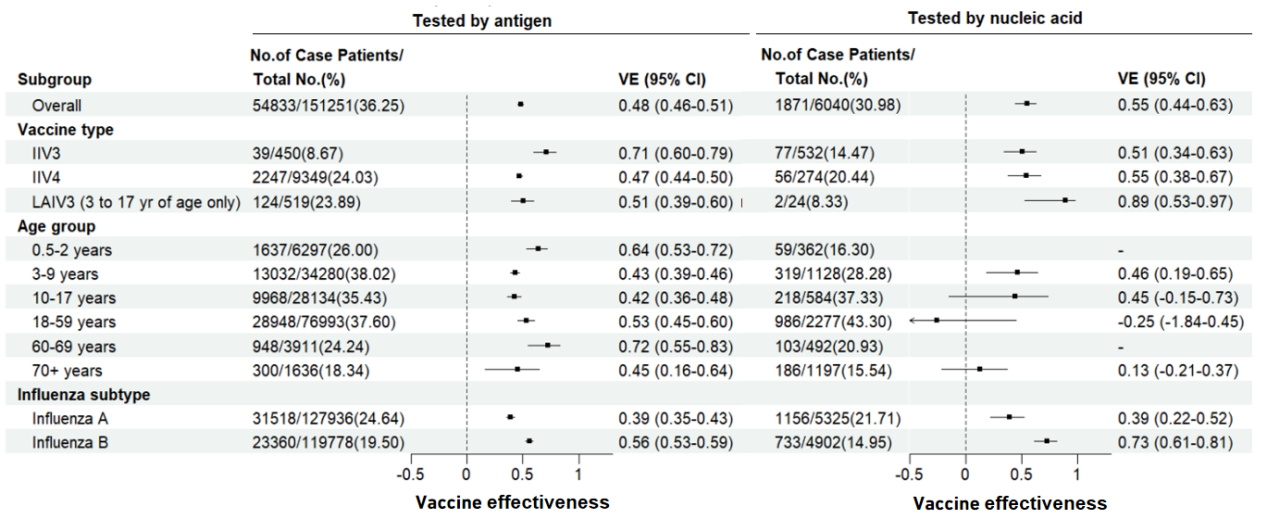

**Figure S3**. Adjusted VE against hospital-attended influenza infection based on antigen and nucleic acid detection results respectively, in the general population and subgroups.

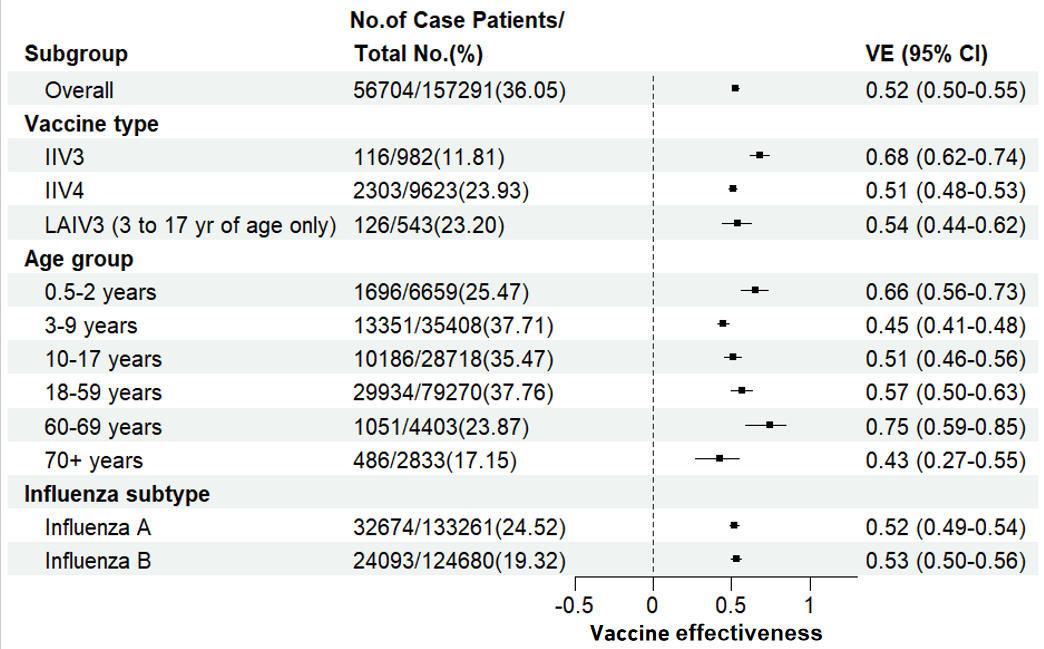

(A)

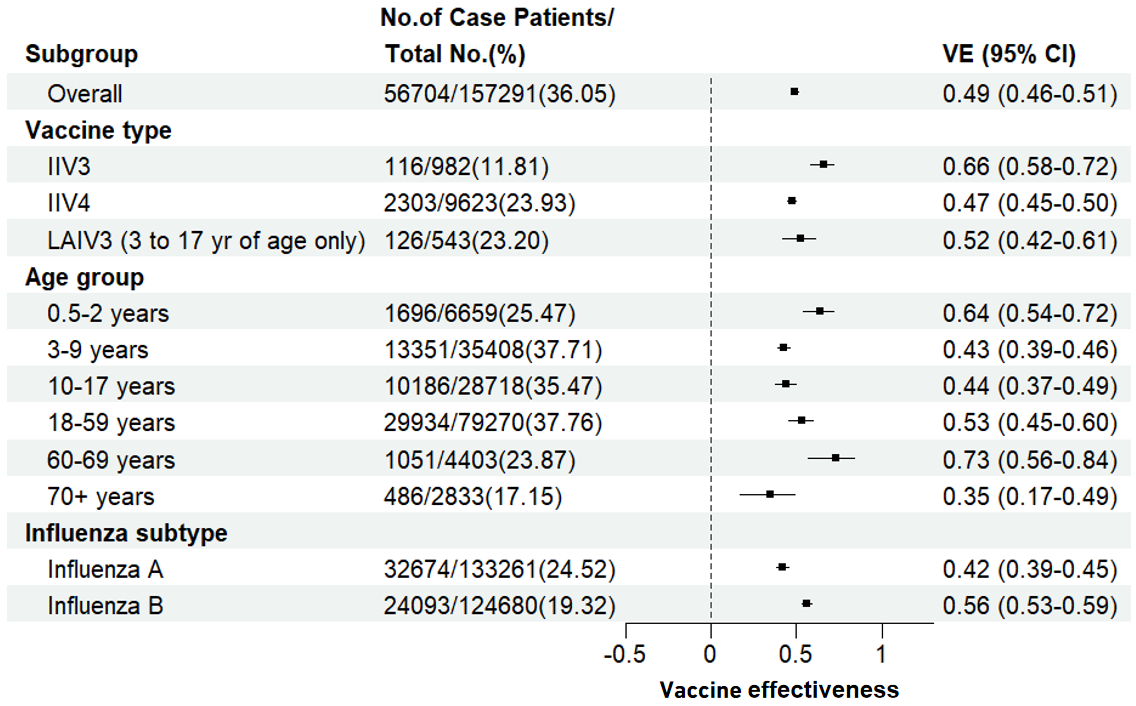

(B)

**Figure S4**. VE against hospital-attended influenza infection (A) without adjusting the influenza testing timing or (B) adjusted by the influenza testing date.

**Supplementary** **Table S1**. Demographic and clinical characteristics of testing-positive and -negative patients for influenza virus based on two influenza detection methods individually.

| Characteristics | Test-positive participants | | | Test-negative participants | Total |
| --- | --- | --- | --- | --- | --- |
|  | Influenza A | Influenza B | A/B coinfection |  |  |
| Tested by antigen | | | | | |
| Total, n (%) | 31473 | 23315 | 45 | 96418 | 151251 |
| Sex, n (%) | | | | | |
| Male | 14848 (47.2%) | 11313 (48.5%) | 20 (44.4%) | 43930 (45.6%) | 70111 (46.4%) |
| Female | 16625 (52.8%) | 12002 (51.5%) | 25 (55.6%) | 52488 (54.4%) | 81140 (53.6%) |
| Age group, n (%) | | | | | |
| 0.5-2 years | 989 (3.1%) | 648 (2.8%) | 0 (0.0%) | 4660 (4.8%) | 6297 (4.2%) |
| 3-9 years | 5579 (17.7%) | 4381 (18.8%) | 8 (17.8%) | 18166 (18.8%) | 28134 (18.6%) |
| 10-17 years | 17097 (54.3%) | 11833 (50.8%) | 18 (40.0%) | 48045 (49.8%) | 76993 (50.9%) |
| 18-59 years | 6957 (22.1%) | 6056 (26.0%) | 19 (42.2%) | 21248 (22.0%) | 34280 (22.7%) |
| 60-69 years | 615 (2.0%) | 333 (1.4%) | 0 (0.0%) | 2963 (3.1%) | 3911 (2.6%) |
| 70+ years | 236 (0.7%) | 64 (0.3%) | 0 (0.0%) | 1336 (1.4%) | 1636 (1.1%) |
| Vaccinated in the 2023/24 season, n (%) | | | | | |
| No | 30101 (95.6%) | 22283 (95.6%) | 39 (86.7%) | 88510 (91.8%) | 140933 (93.2%) |
| Yes | 1372 (4.4%) | 1032 (4.4%) | 6 (13.3%) | 7908 (8.2%) | 10318 (6.8%) |
| Types of Vaccines, n (%) | | | | | |
| IIV3 | 32 (2.3%) | 7 (0.7%) | 0 (0.0%) | 411 (5.2%) | 450 (4.4%) |
| IIV4 | 1242 (90.5%) | 1000 (96.9%) | 5 (83.3%) | 7102 (89.8%) | 9349 (90.6%) |
| LAIV3 | 98 (7.1%) | 25 (2.4%) | 1 (16.7%) | 395 (5.0%) | 519 (5.0%) |
| Tested by nucleic acid | | | | | |
| Total, n (%) | 1138 | 715 | 18 | 4169 | 6040 |
| Sex, n (%) | | | | | |
| Male | 538 (47.3%) | 311 (43.5%) | 9 (50.0%) | 2000 (48.0%) | 2858 (47.3%) |
| Female | 600 (52.7%) | 404 (56.5%) | 9 (50.0%) | 2169 (52.0%) | 3182 (52.7%) |
| Age group, n (%) | | | | | |
| 0.5-2 years | 27 (2.4%) | 31 (4.3%) | 1 (5.6%) | 303 (7.3%) | 362 (6.0%) |
| 3-9 years | 139 (12.2%) | 78 (10.9%) | 1 (5.6%) | 366 (8.8%) | 584 (9.7%) |
| 10-17 years | 584 (51.3%) | 395 (55.2%) | 7 (38.9%) | 1291 (31.0%) | 2277 (37.7%) |
| 18-59 years | 181 (15.9%) | 134 (18.7%) | 4 (22.2%) | 809 (19.4%) | 1128 (18.7%) |
| 60-69 years | 65 (5.7%) | 37 (5.2%) | 1 (5.6%) | 389 (9.3%) | 492 (8.1%) |
| 70+ years | 142 (12.5%) | 40 (5.6%) | 4 (22.2%) | 1011 (24.3%) | 1197 (19.8%) |
| Vaccinated in the 2023/24 season, n (%) | | | | | |
| No | 1037 (91.1%) | 682 (95.4%) | 17 (94.4%) | 3474 (83.3%) | 5210 (86.3%) |
| Yes | 101 (8.9%) | 33 (4.6%) | 1 (5.6%) | 695 (16.7%) | 830 (13.7%) |
| Types of Vaccines, n (%) | | | | | |
| IIV3 | 64 (63.4%) | 12 (36.4%) | 1 (100.0%) | 455 (65.5%) | 532 (64.1%) |
| IIV4 | 36 (35.6%) | 20 (60.6%) | 0 (0.0%) | 218 (31.4%) | 274 (33.0%) |
| LAIV3 | 1 (1.0%) | 1 (3.0%) | 0 (0.0%) | 22 (3.2%) | 24 (2.9%) |

**Supplementary Table S2**. Vaccine coverage in antigen-tested and nucleic acid-tested participants of different age groups.

|  | **Influenza vaccine coverage** | |
| --- | --- | --- |
| **Age** | Tested by antigen | Tested by nucleic acid |
| 0.5-2 years | 665/6297(10.6%) | 25/362(6.9%) |
| 3-9 years | 5941/34280(17.3%) | 176/1128(15.6%) |
| 10-17 years | 2133/28134(7.6%) | 44/584(7.5%) |
| 18-59 years | 1009/76993(1.3%) | 30/2277(1.3%) |
| 60-69 years | 215/3911(5.5%) | 19/492(3.9%) |
| 70+ years | 355/1636(21.7%) | 536/1197(44.8%) |
| **Total** | 10318/151251(6.8%) | 830/6040(13.7%) |

**Supplementary Table S3**. Influenza vaccine coverage in different age groups by vaccine type.

|  | **Influenza vaccine coverage** | | |
| --- | --- | --- | --- |
| **Age** | IIV3 | IIV4 | LAIV3 |
| 0.5-2 years | 75/690(10.9%) | 615/690(89.1%) | 0/690(0.0%) |
| 3-9 years | 27/6117(0.4%) | 5643/6117(92.3%) | 447/6117(7.3%) |
| 10-17 years | 3/2177(0.1%) | 2078/2177(95.5%) | 96/2177(4.4%) |
| 18-59 years | 10/1039(1.0%) | 1029/1039(99.0%) | 0/1039(0.0%) |
| 60-69 years | 18/234(7.7%) | 216/234(92.3%) | 0/234(0.0%) |
| 70+ years | 849/891(95.3%) | 42/891(4.7%) | 0/891(0.0%) |
| **Total** | 982/11148(8.8%) | 9623/11148(86.3%) | 543/11148(4.9%) |
